# Impact of neuropsychiatric symptoms on limbic structures in age-related cognitive decline

**DOI:** 10.1101/2022.06.29.22277055

**Authors:** Ronat Lucas, Hoang Van-Tien, Hanganu Alexandru, the ADNI

**Affiliations:** Centre de Recherche de l’Institut Universitaire de Gériatrie de Montréal, Montréal, Québec, Canada; Faculté de Médecine, Département de Médecine, Université de Montréal, Montréal, Québec, Canada; Faculté des Arts et des Sciences, Département de Psychologie, Université de Montréal, Montréal, Québec, Canada

**Keywords:** Neuropsychiatric symptoms, MRI, Limbic structures, Alzheimer’s Disease, Mild Cognitive Impairment

## Abstract

**Objectives:** Neuropsychiatric symptoms (NPS) are known to increase the risk of cognitive decline in aging. Several studies have investigated the brain substrates of these symptoms, reporting a broad involvement of the limbic regions.

**Participants:** Using the Alzheimer’s Disease Neuroimaging Initiative database, we investigated 223 participants with normal cognition, 367 with mild cognitive impairment (MCI) and 175 with Alzheimer’s disease (AD).

**Measurements:** Neuropsychiatric symptoms were assessed with the Neuropsychiatric Inventory and MRIs were processed with FreeSurfer. We used a general linear model (FreeSurfer) and multivariate analysis of covariance (IBM SPSS software) to establish the associations between the severity of neuropsychiatric symptoms and morphometry in cortical and subcortical structures in each clinical group.

**Results:** The results outlined significant associations between cortical and subcortical structures and neuropsychiatric symptoms. In cognitively normal participants, only a positive association between nighttime behaviors and bilateral caudate nuclei volumes was found. In patients with MCI, agitation, depression, apathy and nighttime behaviors were respectively negatively associated with (i) left precentral and inferior frontal and inferior parietal volumes; (ii) left fusiform volume and area; (iii) right precentral thickness, left frontopolar area and bilateral ventral diencephalon volumes; (iv) right lingual thickness, whereas depression and nighttime behaviors were also respectively positively associated with right ventral diencephalon volume; and left temporal volume and area. In patients with Alzheimer’s disease, broader association were outlined between NPS severity and cortical structures notably agitation, apathy, irritability and nighttime behaviors outlined respectively positive associations with : (i) volumes in the right temporal regions and with surface area in the frontal region; (ii) the cortical thickness of the right pericalcarine region; (iii) volumes in the frontal, temporal and parietal regions; (iv) volume of the right cuneus region; whereas depression and apathy were also respectively negatively associated with the cortical thickness of the left parietal superior region; and cortical volume and area of the parietal regions.

**Conclusions:** These results showed that NPS have broad association patterns with associative brain structures and few associations with limbic structures. These associations were also dependent on the clinical stage of cognitive impairment.

## Introduction

Neuropsychiatric symptoms (NPS), defined as non-cognitive behavioral or mood disturbances.^1,2^ NPS has been shown to precede the dementia stage related to Alzheimer’s Disease (AD) as well as the mild cognitive impairment stage (MCI).^3–5^ Usually, NPS are quantified with the Neuropsychiatric Inventory questionnaire, which measures 12 symptoms and the most common of them are: agitation, depression, anxiety, apathy, irritability, and nighttime behaviors disturbances.^6–8^ Their presence was shown to increase the risk of cognitive decline.^5,9^ They have been associated with structural changes in the limbic system.^10–12^ Specifically, the orbitofrontal, cingulate, subcallosal, insula, temporal lateral and medial, amygdala and thalamus.^13,14^

Overall, the limbic system is involved in emotions and motivation^32^, but region-based association with emotions is different. Some limbic structures appear to be involved in almost all NPS, e.g, cingulate gyrus^10,15–21^, insula^10,18,22–25^ and amygdala^10,26–28^. In the case of the cingulate structures (especially anterior), atrophies could be found in association with agitation^10^, depression^15–17^, anxiety^18^ and apathy^19–21^, whereas hypertrophy was found in cognitively healthy individuals with nighttime behaviors disorders^28^. Data on the insula showed very similar results, with atrophy found in individuals with symptoms of agitation^10,24,25^, anxiety^18^, apathy^18,22,23^ and irritability^23^. If these two structures seem to be co-involved in similar symptoms, they would also underlie different affective systems: cingulate atrophy being involved in depression^15–17^ whereas insular atrophy seems to be involved in irritability^23^. Similarly, to both the cingulate and insular regions, atrophy of the amygdala has been found in agitation^10^, and hypertrophies in anxiety^26^, irritability^27^, and nighttime behaviors disorders^28^. While medial temporal structures such as the hippocampus and parahippocampal gyrus seem to be more specific for agitation^10^ and anxiety^29^ on the one hand and depression and anxiety on the other^10^. Finally, the medial temporal structures, the hippocampus and parahippocampal gyrus, are respectively involved in agitation^5^ and anxiety^11^ on the one hand, and depression and anxiety on the other^18^. This distribution of structures involved in sometimes common NPS raises the question of identical processes underlying entire patterns of NPS. Agitation and anxiety could be linked to almost all the structures elicited. A major literature review by Chen et al. (2021) highlighted numerous limbic, cortical, and subcortical implications in key neuropsychiatric symptoms developed during the pre-dementia stages of AD (including MCI) such as agitation, depression, anxiety, and apathy, as well as other associations with cortical and subcortical associative regions.^30^

Taken together, these data show that some structures appear to be involved in almost all NPS, such as the cingulate and insula, while other structures are more specific to certain symptoms, such as the hippocampus. Interestingly, most studies of NPS have studied participants with a pre-dementia cognitive state (MCI). Few have investigated the brain relationships of NPS in cognitively normal (CN) participants.^21,28,29^ This study proposes to determine cortical and subcortical similarities and divergences between NPS in several stages of cognitive decline (CN, MCI, AD). Also, it is expected that NPS are predominantly characterized by limbic structures.

## Materials and methods

### Participants

Data used in the preparation of this article were obtained from the ADNI database (adni.loni.usc.edu). The ADNI, launched in 2003 and led by Principal Investigator Michael W. Weiner, MD. ADNI aims to combine imaging data (MRI, PET), biological markers, and clinical and neuropsychological assessments to measure the progression of MCI and early AD. ADNI includes 819 adult participants, 55 to 90 years old, who meet entry criteria for a clinical diagnosis of amnestic MCI (n = 397), probable AD (n = 193), or normal cognition (n = 229). Participants received neurological, biological, and neuropsychological assessments at baseline and follow-up visits. (for reviews and more details, see Shaw and colleagues, Mueller and coauthors, and http://www.adni-info.org/)^31–33^. All patients with AD met National Institute of Neurological and Communication Disorders/Alzheimer’s Disease and Related Disorders Association criteria for probable AD with a Mini-Mental State Examination score between 20 and 26, a global Clinical Dementia Rating of 0.5 or 1, a sum-of-boxes Clinical Dementia Rating of 1.0 to 9.0, and, therefore, are only mildly impaired. Entry criteria for patients with amnestic MCI include a Mini-Mental State Examination score of 24 to 30 and a Memory Box score of at least 0.5, whereas other details on the ADNI cohort can be found online.

Thus, 765 participants from the ADNI database were extracted (CN = 223, MCI = 367, AD = 175). Each participant benefited from neuropsychiatric assessment via the Neuropsychiatric Inventory. Six of NPS from the inventory were extracted and analyzed: (i) agitation/aggressiveness, (ii) depression, (iii) anxiety, (iv) apathy, (v) irritability, and (vi) nighttime behavior disorders. Our statistical model included 4 severity values for each of the 6 NPS: no-NPS (a score of zero in the NPI scale), mild NPS (producing little distress in the patient), moderate NPS (more disturbing to the patient but can be redirected by the caregiver) and severe NPS (very disturbing to the patient and difficult to redirect).

Excluding criteria were: (i) incomplete assessments, (ii) incomplete neuropsychiatric and neuropsychological assessments, (iii) presence of psychiatric history (major depression, schizophrenia, bipolar disorder, substance abuse, post-traumatic stress, obsessive-compulsive disorder), (iv) presence of neurological history (stroke, head injury, brain tumor, anoxia, epilepsy, alcohol dependence and Korsakoff, neurodevelopmental disorder), (v) prematurity, (vi) diagnostic criteria in favor of other neurodegenerative or neurological etiology (Parkinson’s disease, frontotemporal degeneration, progressive supranuclear paralysis, corticobasal degeneration, Lewy body dementia, amyotrophic lateral sclerosis, multiple sclerosis, multi-system atrophy, vascular dementia), (vii) use of psychoactive medication (e.g., antidepressants, neuroleptics, chronic anxiolytics, or sedative hypnotics)

Data collection and sharing for this project was funded by the Alzheimer’s Disease Neuroimaging Initiative (ADNI) (National Institutes of Health Grant U01 AG024904) and DOD ADNI (Department of Defense award number W81XWH-12-2-0012). Ethics committee approval and individual patient consents were received by the corresponding registration sites according to ADNI rules (http://adni.loni.usc.edu/methods/documents/). All data are available on the ADNI websites upon demand (http://adni.loni.usc.edu/data-samples/access-data/).

### Data processing and statistical analysis

3T MRI scans were processed on ComputeCanada cluster Cedar with FreeSurfer 7.1.1 software on Linux Centos 7 and managed with an in-house pipeline (github.com/alexhanganu/nimb) that allowed automated exclusion of post-processed data with errors as well as extraction of statistical data, diminishing potential human error. The volumes of subcortical structures were corrected by division with the estimated Total Intracranial Volume.^34^

To assess the effect of each NPS on brain morphology within each clinical group, we performed a whole-brain general linear model analysis with FreeSurfer mri_glmfit for each NPS, based on the Desikan cortical atlas, that includes 34 regions.^35^ Analysis were performed for three cortical measurements: volume, thickness and area. Results underwent a Monte-Carlo correction with a vertex-level threshold of p<0.05.

In a second step, a MANCOVA was performed with IBM SPSS statistics version 26 software to analyze the effect of NPS severity on volumes of subcortical nuclei, within each clinical group (CN, MCI, or AD). The nuclei included in the analysis were: caudate, putamen, pallidum, thalami, nucleus accumbens, ventral diencephalon, hippocampi, and amygdala.^36–38^ NPS severity was defined based on 4 groups: no-NPS, mild NPS, moderate NPS and severe NPS and the comparison contrasts were made between these 4 groups. Post-hoc tests with statistical Tukey correction were performed in the groups for which comparisons were possible due to the size of the subgroups (N > 2), with a threshold of 0.05. Some NPS subgroups could not be considered due to the absence of participants (CN: All-NPS-severe and apathy-moderate; AD: depression-severe). Post-hoc contrasts were performed with a Bonferroni correction (dividing the threshold by the 6, two-by-two inter-group comparisons to be performed for the 4 types of NPS severity: no-NPS vs. mild; no-NPS vs. moderate; no-NPS vs. severe; mild vs. moderate; mild vs. severe; moderate vs. severe). For each post-hoc contrast, the delta value (Δ) indicates the direction of the difference between the groups: a positive value indicates a reduction due to NPS severity; a negative value indicates an increase due to NPS severity.

## Results

Clinical groups consisted of CN participants (mean age: 75.88 ± 5.12), patients with MCI (mean age: 74.83 ± 7.45) and AD (mean age: 75.46 ± 7.47) (Table 1). Groups were similar according to age. The MCI group included more males (63.7%). The mean MMSE score was higher in the CN group compared to MCI and higher in MCI compared to the AD group.

**Table 1:**
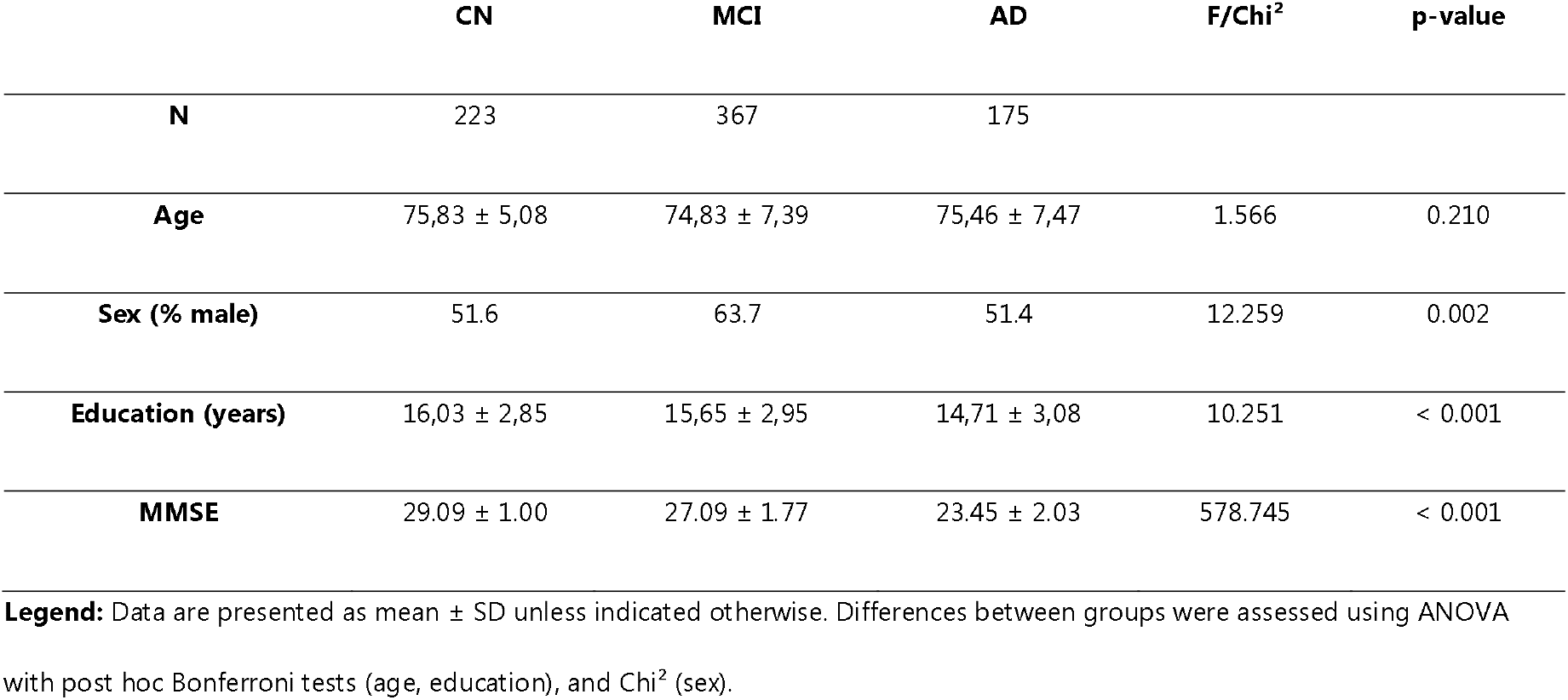
Demographic characteristics according to diagnostic group

### Whole-brain General Linear Model analysis

Monte-Carlo corrected results (Table 2, figure 1) showed that the agitation, depression, apathy and nocturnal behaviors severities were associated with brain structures in the MCI and AD groups. Irritability severity also showed associations but only in the AD group. No association was found in the CN group, nor about anxiety regardless of the group.

**Table 2.**
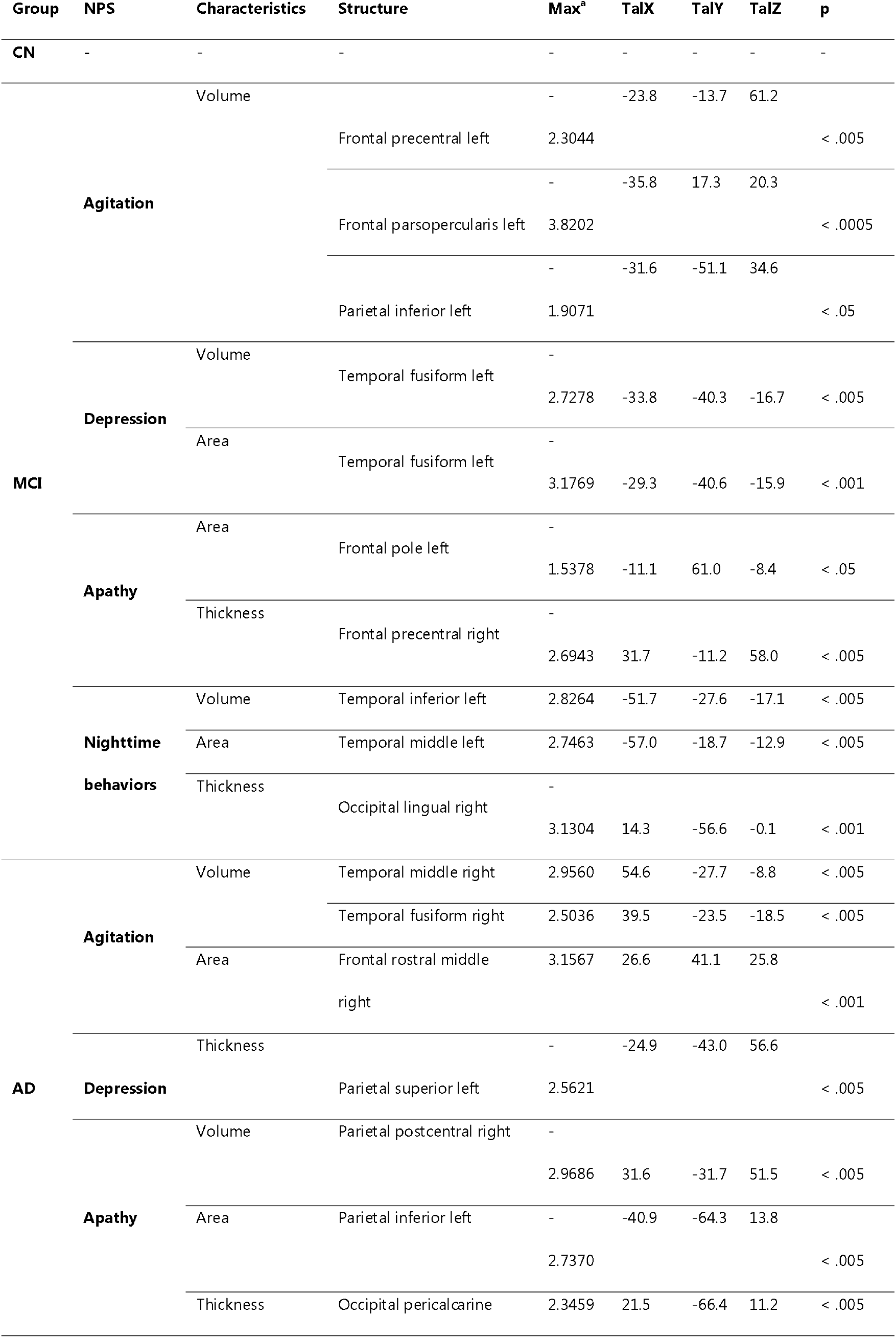
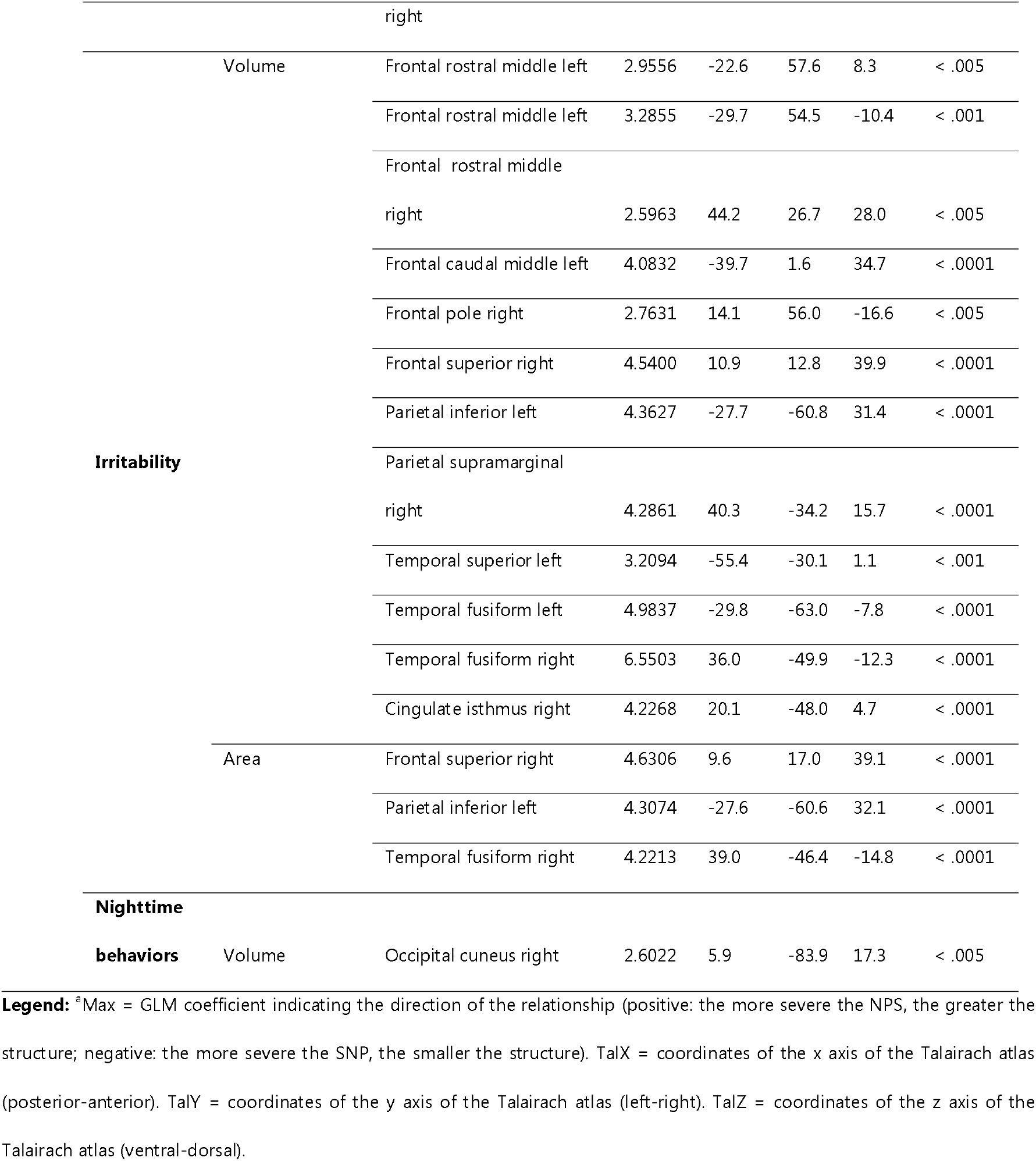
General linear model analysis of NPS severity impacts on brain structures

**Figure 1.**
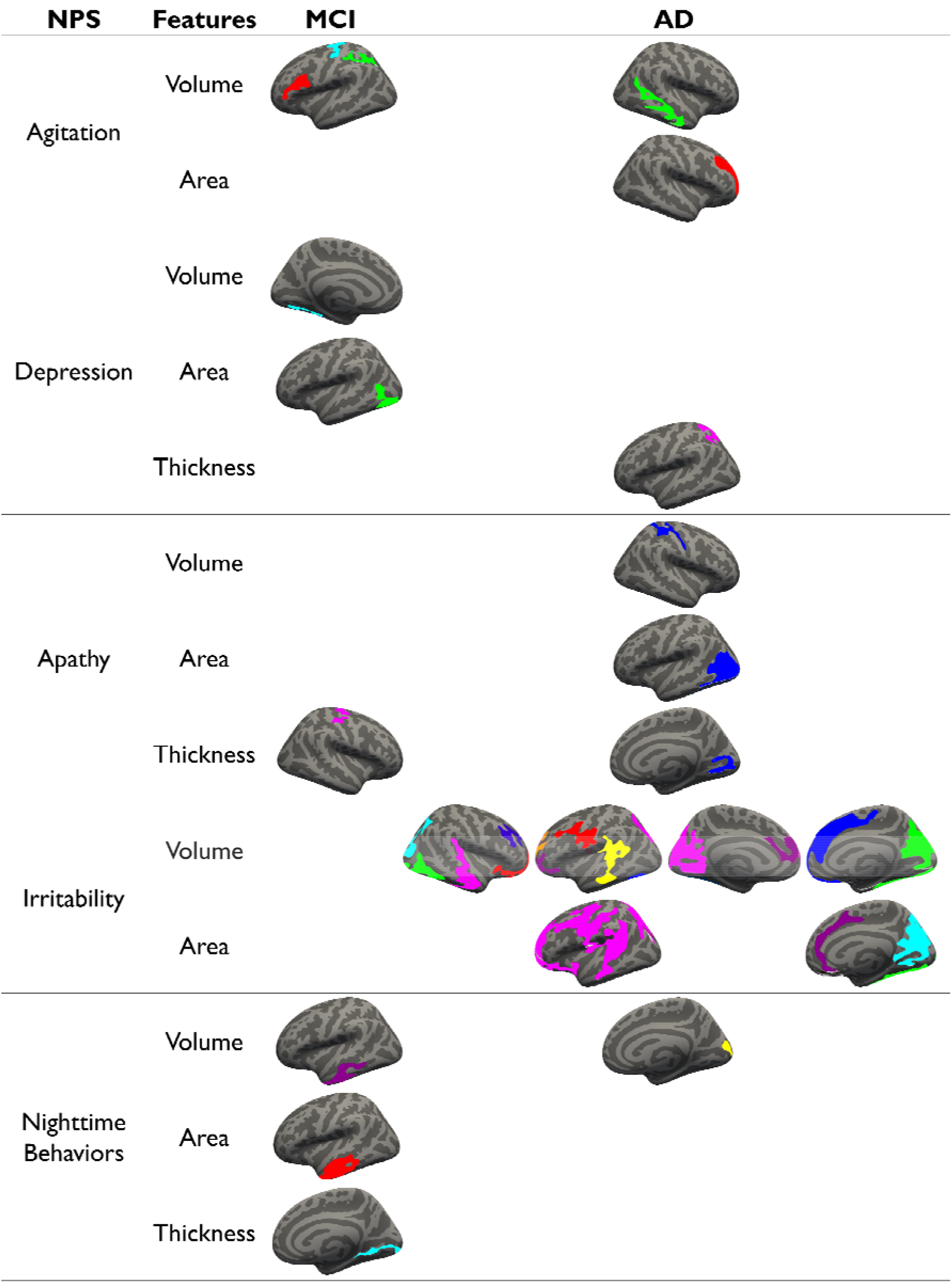
Cortical changes in relation to NPS severity

In the MCI group, agitation severity was negatively associated with left frontal (precentral and *pars opercularis*) and parietal (inferior) volumes. Depression severity was negatively associated with left fusiform volume and area. Apathy severity was negatively associated with right precentral thickness and left frontopolar area. Finally, nighttime behaviors were positively associated with left inferior temporal volume and area, and negatively associated with right lingual thickness.

In the AD group, agitation severity was positively associated with volume changes in the right temporal (middle and fusiform) region and with surface area changes in the frontal (middle) region. Regarding depression, a negative association was found between this NPS and the cortical thickness of the left parietal superior region. Apathy severity was negatively associated with cortical volume and area of the parietal regions (right postcentral and left inferior, respectively) and positively associated with the cortical thickness of the right pericalcarine region. Irritability severity was positively associated with volume changes in the frontal (bilateral middle, left superior and left pole regions), temporal (left fusiform and left superior) and parietal (supramarginal) regions. Some of these volumetric changes were possibly driven by changes in the surface area of the right frontal superior, temporal fusiform and left parietal inferior regions. Nighttime behavior severity was positively associated with volume change of the right cuneus region.

Interestingly, agitation (in MCI and AD) and irritability (in AD) were both associated with temporal (fusiform) and frontal (middle, inferior) structures, whereas depression and apathy in AD were associated with parietal (superior and inferior respectively) structures.

### Univariate subcortical ROI analysis

We also implemented MANCOVA models to investigate the relationships between NPS severity and the volume of subcortical nuclei (Table 3). Post-hoc comparisons showed that nighttime behaviors had an association only in the MCI group whereas depression and apathy severity had an association only in the MCI group. No association in the AD group was found. Finally, agitation, anxiety, and irritability had no significant association in any group.

**Table 3.**
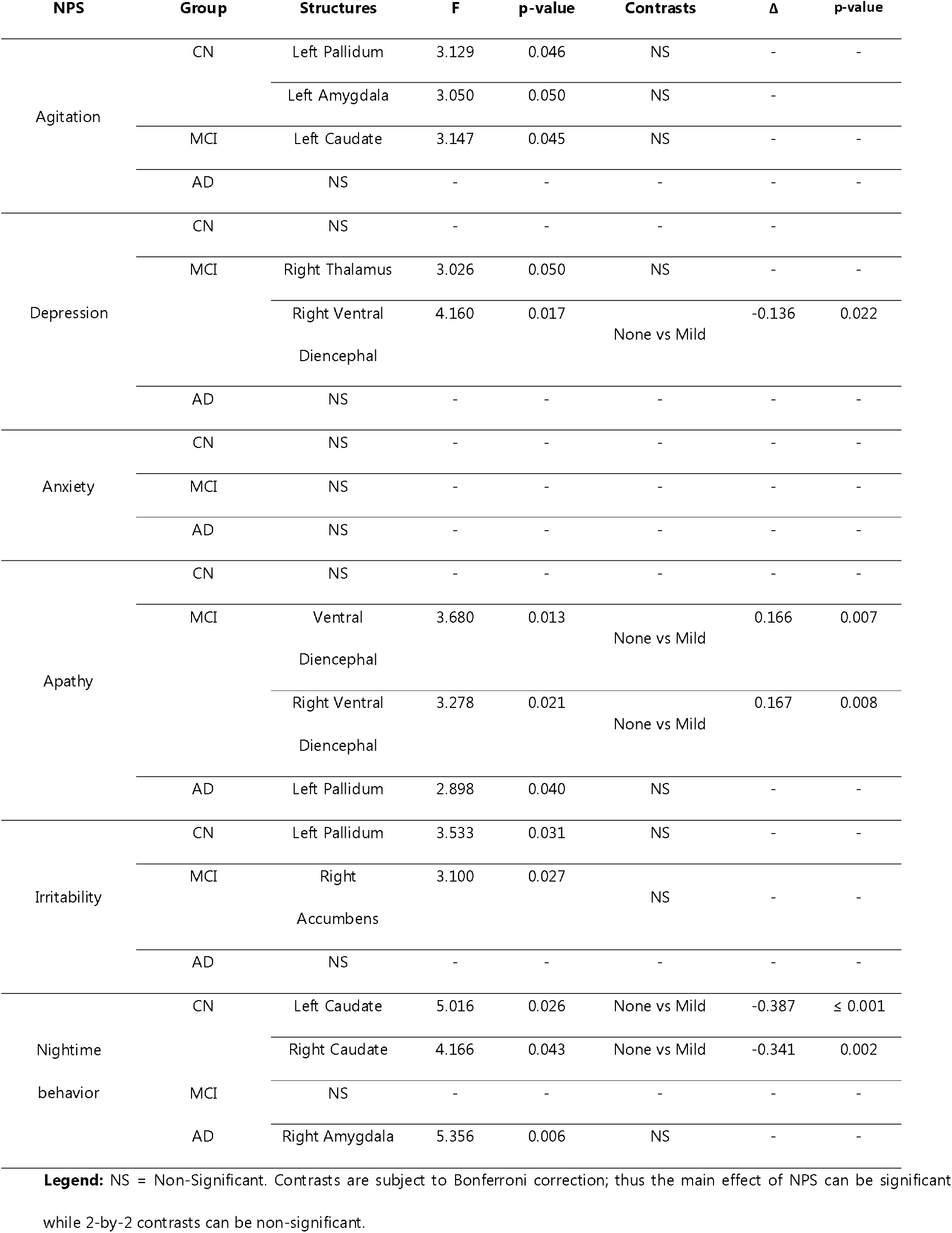
Post-hoc contrasts on the effect of NPS on subcortical brain volumes

In CN participants, only the associations between severity of nighttime behaviors and bilateral caudate nuclei were confirmed by post hoc contrasts, showing that participants with mild nocturnal behavioral disturbances had larger caudate nuclei than those without behavioral disturbances.

In participants with MCI, depression severity was associated with the right ventral diencephalon: participants with mild depression showing a larger structure than those without depression. Conversely, apathy severity was also associated with bilateral ventral diencephalon, but participants with mild apathy showed smaller volumes than those without apathy.

## Discussion

NPS are increasingly known to be involved in age-related cognitive decline. They can anticipate decline and accelerate it.^5,7,9^ Although many studies have looked at their neural correlates, the data diverge and NPS seem to have common and different underlying structures. The objective of this study was to characterize the brain differences associated with NPS severity in groups of different severity of cognitive decline. The most prevalent NPS were studied.

Our results show that: (1) in CN, more severe nighttime behaviors were associated with larger bilateral caudate volumes. No cortical association were found; (2) in MCI, more severe agitation was associated with smaller frontal and parietal volumes; more severe depression was associated with smaller left fusiform volume and area and larger right ventral diencephalon volume; more severe apathy was associated with smaller left frontopolar area, right precentral thickness and ventral bilateral diencephalic volume; more severe nighttime behaviors were associated with smaller left temporal volume and area and greater right lingual thickness; (2a) furthermore, in MCI our results show distinct patterns between depression and nighttime behaviors, respectively negatively and positively associated with the fusiform and inferior left temporal regions; (3) in the AD group, more severe agitation was associated with greater right temporal volume and middle frontal area; more severe depression was associated with smaller right parietal thickness; more severe apathy was associated with smaller right parietal volume and left parietal area, and greater right occipital thickness; more severe irritability was associated with larger frontal, parietal, temporal volumes and areas and cingulate volume; more severe nighttime behaviors were associated with greater right cuneus volume. No subcortical association were found; (3a) finally, in the AD group, we found similarities between depression and apathy, negatively associated with the left parietal region, an opposition with irritability where this region is positively associated, and similarities between agitation and irritability, positively associated with the bilateral temporal and right rostral frontal regions.

These results seem to show a distinct pattern in comparison to previous studies that showed mainly associations with limbic structures.^30^

### Agitation

It was expected that the agitation would be characterized by atrophies in the cingulate, inferior frontal and orbital, insular, amygdala, and hippocampus regions.^10,30,39^ For all the NPS studied, the impacts on brain structures differed from one clinical group to another. Agitation was mainly characterized in CN participants by cortical thinning of the lateral temporal regions and right medial subcallosal and orbitofrontal thickening, whereas in MCI patients, agitation was more related to bilateral insular thinning and an increase in the volume of the left central amygdala. In AD patients, agitation was subtended by insular, temporo-polar, left anterior thalamic and right thalamic thinning. The GLM analysis showed the effect of agitation only in the AD group by thickening the right superior and middle rostral frontal gyri. Overall, these data are consistent with the previous literature concerning the amygdala, orbitofrontal regions, insular and cingulate regions.^10,24,25^ Indeed, the literature showed multiple implications in agitation such as atrophies of anterior structures like cingular, frontal, insular, amygdala and hippocampal atrophies and posterior structures like medial and posterior parietal, cingular and frontal cortices in MCI and AD populations.^10,24,25^ But no one showed relations with temporal or thalamic structures.

### Depression

According to the literature, numerous studies have shown the diversity of the regions impacted during the presence of depression linked to ageing and in particular AD, more particularly atrophies and volume reductions which can affect the whole brain.^40^ More recently, from the literature review by Chen et al. (2021), depression is associated with extensive lesions, suggesting the involvement of extensive networks. These lesions involve frontal (superior, medial, dorsolateral), cingulate (anterior and posterior), parietal (inferior, supramarginal, precuneus), temporal (superior and inferior, entorhinal, fusiform, hippocampus), and subcortical (caudate) regions. These data suggest damage to the fronto-limbic network.^30^ Our results go against these data. Indeed, according to the present analyses, Depression severity was only negatively associated with superior frontal gyrus thickness in CN subjects by the general linear model. No relationship was found in the other groups. This lack of difference contrasts with most of the results reported in the previous literature. Depression is probably the NPS the more studied in AD and cognitive decline. Also, many studies demonstrated a lot of structures with greater atrophy in association with depression in MCI and AD. These structures concerned medial temporal lobe, temporopolar, entorhinal, parahippocampal, cingular and middle frontal cortices.^6,9,10,34,36^ Association between depression and apathy and their impacts on the atrophy pathways of MCI participants was studied by Zahodne *et al*.^16^ Their results showed that the associations were not significant. Thus, depression was estimated as a better marker of cortical atrophy.^16^ However, the results from the current study showed only impact of depression in the CN group with the GLM analysis and only in CN and MCI groups by the ANOVA. So, it matches with the explanation from Lee et al. who suggested that cerebral and cognitive changes associated with AD may obscure changes associated with subsyndromal depression.^41^ These changes associated with neuropsychiatric depression could also be obscured by the impact of the physiopathological process of AD.

### Anxiety

Anxiety would be characterized by hippocampal, insular, parahippocampal, and cingulate limbic atrophies, as well as amygdala hypertrophy.^18,26,29,42^ Also, anxiety was mainly associated with entorhinal, parietal, temporal, frontal, and putamen lesions.^30^ Amygdala hypertrophy is indeed found in CN participants, particularly on the left in the anterior amygdaloid area. The GLM analysis found others increase of cortical areas for left posterior cingulate and precuneus. Cingular thinning was found in MCI patients, particularly on the left in the posterior ventral region. In AD patients, the implications are more extensive and concern right parahippocampal, subcallosal and temporal cortical thinning, as well as reductions in right, lateral medial and medial thalamic volumes. The literature showed more posterior and subcortical atrophies essentially in amnesic MCI and AD. These atrophies concerned temporal medial regions, insula, cingulate posterior and putamen.^18,42^ Moreover, it was also demonstrated to increase volumes of amygdala and caudate nucleus volume.^26^ Thus, these data confirm the previous literature. However, if the structures involved are related to this literature, they remain very diverse according to the clinical stages in the sense that the clinical severity is characterized by a greater number of structures involved, which runs counter to certain studies which have demonstrated the impact of anxiety in CN participants but not in MCI or AD patients.

### Apathy

Based on the literature review by Chen et al. (2021), apathy is associated with predominantly left lesions of: anterior cingulate, medial prefrontal orbitofrontal, superior frontal, parietal, and caudate nuclei and putamen.^30^ Also, apathy would rather concern the posterior cingulate gyrus (MCI and AD), the inferior frontal and medial, insular, and inferior temporal (CN) regions.^18–20,22,23,39^ In CN participants, only the insular thinning (left) is found. Similarly, in MCI patients, only bilateral anterior thalamic thinning is found. Left insular thinning is also found in AD patients, as well as a thickening of the callosomarginal sulcus. The GLM analysis highlighted association between the severity of apathy and left parahippocampal thickening and postcentral gyrus thinning in MCI group, whereas there were associations with decrease of left inferior parietal area and right pericalcarine sulcus thickening. According to the data in the literature, apathy is involved in many structures, which the current study does not corroborate. Furthermore, it is necessary to specify that there are different types of apathy, being involved by atrophies of various subcortical-frontal loops.^43^ Most studies do not specify the type of apathy being studied, particularly because the assessment scales are not always designed to distinguish between them. Furthermore, the evaluation of apathy does not always allow for verification of the process that causes it (cognitive, affective, or motivational). Some studies have shown a relationship with inferior temporal regions or previous thalamic radiation with apathy.^44,45^ One could suppose that these regions, respectively involved in language and memory processes, would induce the loss of associated functions when affected by neurodegeneration processes, and would make individuals less able to communicate, leading to isolation, withdrawal and may be interpreted as apathy by relatives. The fact that there is so much different data in the literature on the cerebral substrates of apathy shows the complexity of this notion and the processes that underlie it.

### Irritability

Irritability, probably one of the least studied NPS, would be linked to bilateral insular atrophies, notably anterior, but also posterior right insular atrophy in the patients, decreased white matter integrity in the anterior cingulate cortex, as well as amygdala preservation in AD participants.^23,27,46^ The available data on the cerebral substrates of irritability are poor in the literature. The same was true of its impact on cognition. So, our results complement the available data and showed that irritability is characterized by a left temporo-polar thinning and a right inferior temporal thickening. In MCI patients, irritability is related to a left anterior insular thinning, whereas in AD patients, irritability is characterized by an increase in volume of the left central amygdala, which agrees with some of the data in the literature. From the literature, only reduced volume of right insula and preserve volume of amygdala in AD was showed.^23,27^

### Nighttime behaviors

Finally, nighttime behaviors would be characterized by increases in brain volume in the frontal limbic, temporal and amygdala regions, and by atrophies in participants in similar regions according to the progress of the clinical stages.^28,47,48^ These data are particularly derived from work on the cerebral impacts of obstructive sleep apnea, generating up to hundreds of micro-awakenings per night, disrupting the paradoxical and general sleep of hypoxia.^28,47^ The results of the study confirm previous data, in particular: in the CN group, in which nighttime behaviors are characterized by inferior temporal and left orbital-frontal cortical thickenings, as well as increases in amygdala volumes involving the left cortico-amygdaloid and bilateral paralaminar nuclei. Lower temporal thickening was found in MCI patients with right insular and orbital-frontal thinning. Finally, interestingly, the presence of these disorders in AD patients showed insular cortical, parahippocampal, left temporal-polar, bilateral orbitofrontal thinning and increases in the subcortical volumes of the right basal, central, medial and paralaminar amygdala nuclei, the right global amygdala, and the right lateral medial and right global thalamus nucleus. In the case of these sleep disorders, studies including sleep apnea have shown both increases and decreases in the size of cortical and subcortical structures.^20,21^ These increases and decreases are related to inflammatory processes, according to the authors. These processes increase cell size initially, resulting in an increase in cortical thickness, until cell death and therefore degeneration, resulting in reductions in cortical thickness and subcortical volumes in a second stage.^28,47^ This may explain the ANOVA results showing increases in volumes and thicknesses in CN participants, thickening and thinning in MCI patients and cortical thinning and increases in subcortical volumes in AD patients.

### NPS and brain morphology

Some limbic structures appear to be involved in almost all NPS, e.g, cingulate gyrus^10,15–21^, insula^10,18,22–25^ and amygdala^10,26–28^. This could suggest the existence of a common network for some NPS. However, our results also highlighted associations with associative cortical structures (fusiform and middle temporal, inferior parietal, prefrontal regions). These raise the question of the relationship between NPS and cognitive processes underpinned by prefrontal, temporal and parietal regions such as executive, language and communicative functions. Also, it cannot be excluded that some NPS, particularly in participants with MCI or AD, are consecutive to cognitive disorders (apathy due to executive difficulties, irritability due to memory difficulties) and to the reduction of functional abilities. Furthermore, the associations with ventral diencephalic regions and caudate nuclei suggest the involvement of social-cognitive rather than emotional processes in NPS.

Analysis of the NPS most frequently found in the samples studied shows both diverse and partially similar neuroanatomical characteristics. Indeed, some NPS present similar cortical and/or subcortical characteristics. This neuroanatomical overlap between some NPS could underlie the identification of NPS clusters.^49^ These characteristics vary according to the clinical and control groups. Interestingly, cerebral differences can even be found in CN participants, which raises the question of the fragility of certain regions and processes related to NPS. These fragilizations could be a privileged route of entry into MCI. As shown by longitudinal studies, some NPS may be present several years before a clinical state of MCI for CN participants, or a state of dementia for MCI patients, emerges.^3,4,9,16^ Therefore, even if the prevalence of NPS is lower in CN participants, they constitute a population of choice for understanding the emergence of neuronal disorders and fragility.

Similarly, to functional connectivity methods, whole-brain studies have several advantages, but such analyses must use stringent statistical corrections to account for multiple comparison issues given the large voxel number. Thus, the statistical threshold at which these studies report their results can have a huge impact on the replicability of the findings. In addition, connectivity results reported at different levels (whole brain and a priori ROIs) can not only help conceptualizing the findings better but also in improving the replicability of findings. Thus, future studies should take the aforementioned factors into account to improve the reliability and replicability of connectivity studies.^50^

### Limitations

Several limitations are to be considered in this study, starting with the transversal aspect of the experimental protocol, which does not allow for a causal relationship between NPS and their cognitive and cerebral characteristics. Moreover, the NPS presents strong comorbidities between them, most of the patients presented several NPS in their neuropsychiatric profile. It would therefore be interesting to study the impact of combinations between NPS rather than NPS in isolation. Also, the study of the severity of NPS induces an increase in intergroup heterogeneity, both in terms of sample sizes (some subgroups may be too small for post-hoc analyses), and in terms of brain characteristics. Thus, some of the differences highlighted could be explained more by the characteristics of the individual rather than by the presence of a particular NPS.

## Conclusion

In conclusion, in a large cohort of the ADNI database, we showed that NPS were related to changes in cortical and subcortical structures mainly to the prefrontal and medial temporal structures on the one hand, as well as extensively in the limbic system on the other hand. The impact of NPS was found in all clinical groups, even in CN participants. However, these impacts differed between groups and between NPS. Notably, changes were due to anxiety in the CN participants, irritability and nighttime behaviors in all groups, agitation and apathy in the groups characterized by cognitive impairment (MCI and AD). These data suggest that some NPS may occur earlier than others while impacting brain structures. Further studies, especially longitudinal ones, should investigate the cognitive trajectories of CN and MCI participants according to the NPS/brain structure relationships they present.

## Data Availability

All data are available on the Alzheimers Disease Neuroimaging Initiative websites upon demand (http://adni.loni.usc.edu/data-samples/access-data/).

## Abbreviations

ADNI: Alzheimer’s Disease Neuroimaging Initiative
CN: Cognitively Normal
GLM: General Linear Model
MMSE: MiniMental State Examination
NPI: Neuropsychiatric Inventory
NPS: Neuropsychiatric Symptoms
WAIS: Wechsler Adult Intelligence Scale.

## AUTHORS’ CONTRIBUTIONS

Research project: AH, LR; Data extraction and processing: VTH, AH, LR; Statistical analysis: LR; Manuscript: LR, AH

## Use of ADNI data

*Data collection and sharing for this project was funded by the Alzheimer’s Disease Neuroimaging Initiative (ADNI) (National Institutes of Health Grant U01 AG024904) and DOD ADNI (Department of Defense award number W81XWH-12-2-0012). ADNI is funded by the National Institute on Aging, the National Institute of Biomedical Imaging and Bioengineering, and through generous contributions from the following: AbbVie, Alzheimer’s Association; Alzheimer’s Drug Discovery Foundation; Araclon Biotech; BioClinica, Inc*.; *Biogen; Bristol-Myers Squibb Company; CereSpir, Inc*.; *Cogstate; Eisai Inc*.; *Elan Pharmaceuticals, Inc*.; *Eli Lilly and Company; EuroImmun; F. Hoffmann-La Roche Ltd and its affiliated company Genentech, Inc*.; *Fujirebio; GE Healthcare; IXICO Ltd*.; *Janssen Alzheimer Immunotherapy Research & Development, LLC*.; *Johnson & Johnson Pharmaceutical Research & Development LLC*.; *Lumosity; Lundbeck; Merck & Co*., *Inc*.; *Meso Scale Diagnostics, LLC*.; *NeuroRx Research; Neurotrack Technologies; Novartis Pharmaceuticals Corporation; Pfizer Inc*.; *Piramal Imaging; Servier; Takeda Pharmaceutical Company; and Transition Therapeutics. The Canadian Institutes of Health Research is providing funds to support ADNI clinical sites in Canada. Private sector contributions are facilitated by the Foundation for the National Institutes of Health (www.fnih.org). The grantee organization is the Northern California Institute for Research and Education, and the study is coordinated by the Alzheimer’s Therapeutic Research Institute at the University of Southern California. ADNI data are disseminated by the Laboratory for NeuroImaging at the University of Southern California*.

## Appendix 1

### ADNI Co-investigator

Michael Weiner, MD (UC San Francisco, PI of ADNI), Paul Aisen, MD (UC San Diego, ADCS PI and Director of Coordinating Center Clinical Core), Michael Weiner, MD (UC San Francisco, Executive Committee), Paul Aisen, MD (UC San Diego, Executive Committee), Ronald Petersen, MD, PhD (Mayo Clinic, Rochester, Executive Committee), Clifford R. Jack, Jr., MD (Mayo Clinic, Rochester, Executive Committee), William Jagust, MD (UC Berkeley, Executive Committee), John Q. Trojanowki, MD, PhD (U Pennsylvania, Executive Committee), Arthur W. Toga, PhD (UCLA, Executive Committee), Laurel Beckett, PhD (UC Davis, Executive Committee), Robert C. Green, MD, MPH (Brigham and Women’s Hospital/Harvard Medical School, Executive Committee), Andrew J. Saykin, PsyD (Indiana University, Executive Committee), John Morris, MD (Washington University St. Louis, Executive Committee), Enchi Liu, PhD (Janssen Alzheimer Immunotherapy, ADNI 2 Private Partner Scientific Board (Chair), Robert C. Green, MD, MPH (Brigham and Women’s Hospital/Harvard Medical School, Data and Publication Committee (Chair)), Tom Montine, MD, PhD (University of Washington, Resource Allocation Review Committee), Ronald Petersen, MD, PhD (Mayo Clinic, Rochester, Clinical Core Leaders (Core PI)), Paul Aisen, MD (UC San Diego, Clinical Core Leaders), Anthony Gamst, PhD (UC San Diego, Clinical Informatics and Operations), Ronald G. Thomas, PhD (UC San Diego, Clinical Informatics and Operations), Michael Donohue, PhD (UC San Diego, Clinical Informatics and Operations), Sarah Walter, MSc (UC San Diego, Clinical Informatics and Operations), Devon Gessert (UC San Diego, Clinical Informatics and Operations), Tamie Sather (UC San Diego, Clinical Informatics and Operations), Laurel Beckett, PhD (UC Davis, Biostatistics Core Leaders and Key Personnel (Core PI)), Danielle Harvey, PhD (UC Davis, Biostatistics Core Leaders and Key Personnel), Anthony Gamst, PhD (UC San Diego, Biostatistics Core Leaders and Key Personnel), Michael Donohue, PhD (UC San Diego, Biostatistics Core Leaders and Key Personnel), John Kornak, PhD (UC Davis, Biostatistics Core Leaders and Key Personnel), Clifford R. Jack, Jr., MD (Mayo Clinic, Rochester, MRI Core Leaders and Key Personnel (Core PI)), Anders Dale, PhD (UC San Diego, MRI Core Leaders and Key Personnel), Matthew Bernstein, PhD (Mayo Clinic, Rochester, MRI Core Leaders and Key Personnel), Joel Felmlee, PhD (Mayo Clinic, Rochester, MRI Core Leaders and Key Personnel), Nick Fox, MD (University of London, MRI Core Leaders and Key Personnel), Paul Thompson, PhD (UCLA School of Medicine, MRI Core Leaders and Key Personnel), Norbert Schuff, PhD (UCSF MRI, MRI Core Leaders and Key Personnel), Gene Alexander, PhD (Banner Alzheimer’s Institute, MRI Core Leaders and Key Personnel), Charles DeCarli, MD (UC Davis, MRI Core Leaders and Key Personnel), William Jagust, MD (UC Berkeley, PET Core Leaders and Key Personnel (Core PI)), Dan Bandy, MS, CNMT (Banner Alzheimer’s Institute, PET Core Leaders and Key Personnel), Robert A. Koeppe, PhD (University of Michigan, PET Core Leaders and Key Personnel), Norm Foster, MD (University of Utah, PET Core Leaders and Key Personnel), Eric M. Reiman, MD (Banner Alzheimer’s Institute, PET Core Leaders and Key Personnel), Kewei Chen, PhD (Banner Alzheimer’s Institute, PET Core Leaders and Key Personnel), Chet Mathis, MD (University of Pittsburgh, PET Core Leaders and Key Personnel), John Morris, MD (Washington University St. Louis, Neuropathology Core Leaders), Nigel J. Cairns, PhD, (MRCPath Washington University St. Louis, Neuropathology Core Leaders), Lisa Taylor-Reinwald, BA, HTL (Washington University St. Louis, Neuropathology Core Leaders), J.Q. Trojanowki, MD, PhD (UPenn School of Medicine, Biomarkers Core Leaders and Key Personnel (Core PI)), Les Shaw, PhD (UPenn School of Medicine, Biomarkers Core Leaders and Key Personnel), Virginia M.Y. Lee, PhD, MBA (UPenn School of Medicine, Biomarkers Core Leaders and Key Personnel), Magdalena Korecka, PhD (UPenn School of Medicine, Biomarkers Core Leaders and Key Personnel), Arthur W. Toga, PhD (UCLA, Informatics Core Leaders and Key Personnel (Core PI)), Karen Crawford (UCLA, Informatics Core Leaders and Key Personnel), Scott Neu, PhD (UCLA, Informatics Core Leaders and Key Personnel), Andrew J. Saykin, PsyD (Indiana University, Genetics Core Leaders and Key Personnel), Tatiana M. Foroud, PhD (Indiana University, Genetics Core Leaders and Key Personnel), Steven Potkin, MD (UC Irvine, Genetics Core Leaders and Key Personnel), Li Shen, PhD (Indiana University, Genetics Core Leaders and Key Personnel), Zaven Kachaturian, PhD (Khachaturian, Radebaugh & Associates (KRA), Inc, Early Project Development), Richard Frank, MD, PhD (General Electric, Early Project Development), Peter J. Snyder, PhD (University of Connecticut, Early Project Development), Susan Molchan, PhD (National Institute on Aging/National Institutes of Health, Early Project Development), Jeffrey Kaye, MD (Oregon Health and Science University, Site Investigator), Joseph Quinn, MD (Oregon Health and Science University, Site Investigator), Betty Lind, BS (Oregon Health and Science University, Site Investigator), Sara Dolen, BS (Oregon Health and Science University, Past Site Investigator), Lon S. Schneider, MD (University of Southern California, Site Investigator), Sonia Pawluczyk, MD (University of Southern California, Site Investigator), Bryan M. Spann, DO, PhD (University of Southern California, Site Investigator), James Brewer, MD, PhD (University of California--San Diego, Site Investigator), Helen Vanderswag, RN (University of California--San Diego, Site Investigator), Judith L. Heidebrink, MD, MS, (University of Michigan, Site Investigator), Joanne L. Lord, LPN, BA, CCRC (University of Michigan, Site Investigator), Ronald Petersen, MD, PhD (Mayo Clinic, Rochester, Site Investigator), Kris Johnson, RN (Mayo Clinic, Rochester, Site Investigator), Rachelle S. Doody, MD, PhD (Baylor College of Medicine, Site Investigator), Javier Villanueva-Meyer, MD (Baylor College of Medicine, Site Investigator), Munir Chowdhury, MBBS, MS (Baylor College of Medicine, Site Investigator), Yaakov Stern, PhD (Columbia University Medical Center, Site Investigator), Lawrence S. Honig, MD, PhD (Columbia University Medical Center, Site Investigator), Karen L. Bell, MD, (Columbia University Medical Center, Site Investigator), John C. Morris, MD (Washington University, St. Louis, Site Investigator), Beau Ances, MD (Washington University, St. Louis, Site Investigator), Maria Carroll, RN, MSN (Washington University, St. Louis, Site Investigator), Sue Leon, RN, MSN (Washington University, St. Louis, Site Investigator), Mark A. Mintun, MD (Washington University, St. Louis, Past Site Investigator), Stacy Schneider, APRN, BC, GNP (Washington University, St. Louis, Past Site Investigator), Daniel Marson, JD, PhD (University of Alabama – Birmingham, Site Investigator), Randall Griffith, PhD, ABPP (University of Alabama – Birmingham, Site Investigator), David Clark, MD (University of Alabama – Birmingham, Site Investigator), Hillel Grossman, MD (Mount Sinai School of Medicine, Site Investigator), Effie Mitsis, PhD (Mount Sinai School of Medicine, Site Investigator), Aliza Romirowsky, BA (Mount Sinai School of Medicine, Site Investigator), Leyla deToledo-Morrell, PhD (Rush University Medical Center, Site Investigator), Raj C. Shah, MD (Rush University Medical Center, Site Investigator) (Wein Center, Site Investigator), Ranjan Duara, MD (Wein Center, Site Investigator), Daniel Varon, MD (Wein Center, Site Investigator), Peggy Roberts, CNA (Wein Center, Site Investigator), Marilyn Albert, PhD (Johns Hopkins University, Site Investigator), Chiadi Onyike, MD, MHS (Johns Hopkins University, Site Investigator), Stephanie Kielb MD (Johns Hopkins University, Site Investigator), Henry Rusinek, PhD (New York University, Site Investigator), Mony J de Leon, EdD (New York University, Site Investigator), Lidia Glodzik, MD, PhD (New York University, Site Investigator), Susan De Santi, PhD (New York University, Past Site Investigator), P. Murali Doraiswamy, MD (Duke University Medical Center, Site Investigator), Jeffrey R. Petrella, MD (Duke University Medical Center, Site Investigator), R. Edward Coleman, MD (Duke University Medical Center, Site Investigator), Steven E. Arnold, MD (University of Pennsylvania, Site Investigator), Jason H. Karlawish, MD, (University of Pennsylvania, Site Investigator), David Wolk, MD (University of Pennsylvania, Site Investigator), Charles D. Smith, MD (University of Kentucky, Site Investigator), Greg Jicha, MD (University of Kentucky, Site Investigator), Peter Hardy, PhD (University of Kentucky, Site Investigator), Oscar L. Lopez, MD (University of Pittsburgh, Site Investigator), MaryAnn Oakley, MA (University of Pittsburgh, Site Investigator), Donna M. Simpson, CRNP, MPH (University of Pittsburgh, Site Investigator), Anton P. Porsteinsson, MD (University of Rochester Medical Center, Site Investigator), Bonnie S. Goldstein, MS, NP (University of Rochester Medical Center, Site Investigator), Kim Martin, RN (University of Rochester Medical Center, Site Investigator), Kelly M. Makino, BS (University of Rochester Medical Center, Past Site Investigator), M. Saleem Ismail, MD (University of Rochester Medical Center, Past Site Investigator), Connie Brand, RN (University of Rochester Medical Center, Past Site Investigator), Ruth A. Mulnard, DNSc, RN, FAAN (University of California, Irvine, Site Investigator), Gaby Thai, MD (University of California, Irvine, Site Investigator), Catherine Mc-Adams-Ortiz, MSN, RN, A/GNP (University of California, Irvine, Site Investigator), Kyle Womack, MD (University of Texas Southwestern Medical School, Site Investigator), Dana Mathews, MD, PhD (University of Texas Southwestern Medical School, Site Investigator), Mary Quiceno, MD (University of Texas Southwestern Medical School, Site Investigator), Ramon Diaz-Arrastia, MD, PhD (University of Texas Southwestern Medical School, Past Site Investigator), Richard King, MD (University of Texas Southwestern Medical School, Past Site Investigator), Myron Weiner, MD (University of Texas Southwestern Medical School, Past Site Investigator), Kristen Martin-Cook, MA (University of Texas Southwestern Medical School, Past Site Investigator), Michael DeVous, PhD (University of Texas Southwestern Medical School, Past Site Investigator), Allan I. Levey, MD, PhD (Emory University, Site Investigator), James J. Lah, MD, PhD (Emory University, Site Investigator), Janet S. Cellar, DNP, PMHCNS-BC (Emory University, Site Investigator), Jeffrey M. Burns, MD (University of Kansas, Medical Center, Site Investigator), Heather S. Anderson, MD (University of Kansas, Medical Center, Site Investigator), Russell H. Swerdlow, MD (University of Kansas, Medical Center, Site Investigator), Liana Apostolova, MD (University of California, Los Angeles, Site Investigator), Po H. Lu, PsyD (University of California, Los Angeles, Site Investigator), George Bartzokis, MD (University of California, Los Angeles, Past Site Investigator), Daniel H.S. Silverman, MD, PhD (University of California, Los Angeles, Past Site Investigator), Neill R Graff-Radford, MBBCH, FRCP (London) (Mayo Clinic, Jacksonville, Site Investigator), Francine Parfitt, MSH, CCRC (Mayo Clinic, Jacksonville, Site Investigator), Heather Johnson, MLS, CCRP (Mayo Clinic, Jacksonville, Site Investigator), Martin R. Farlow, MD (Indiana University, Site Investigator), Ann Marie Hake, MD, Brandy R. Matthews, MD (Indiana University, Site Investigator), Scott Herring, RN (Indiana University, Past Site Investigator), Christopher H. van Dyck, MD (Yale University School of Medicine, Site Investigator), Richard E. Carson, PhD (Yale University School of Medicine, Site Investigator), Martha G. MacAvoy, PhD (Yale University School of Medicine, Site Investigator), Howard Chertkow, MD (McGill Univ., Montreal-Jewish General Hospital, Site Investigator), Howard Bergman, MD (McGill Univ., Montreal-Jewish General Hospital, Site Investigator), Chris Hosein, MEd (McGill Univ., Montreal-Jewish General Hospital, Site Investigator), Sandra Black, MD, FRCPC (Sunnybrook Health Sciences, Ontario, Site Investigator), Bojana Stefanovic, PhD (Sunnybrook Health Sciences, Ontario, Site Investigator), Curtis Caldwell, PhD (Sunnybrook Health Sciences, Ontario, Site Investigator), Ging-Yuek Robin Hsiung, MD, MHSc, FRCPC (U.B.C. Clinic for AD & Related Disorders, Site Investigator), Howard Feldman, MD, FRCPC (U.B.C. Clinic for AD & Related Disorders, Site Investigator), Benita Mudge, BS (U.B.C. Clinic for AD & Related Disorders, Site Investigator), Michele Assaly, MA (U.B.C. Clinic for AD & Related Disorders, Past Site Investigator), Andrew Kertesz, MD (Cognitive Neurology - St. Joseph’s, Ontario, Site Investigator), John Rogers, MD (Cognitive Neurology - St. Joseph’s, Ontario, Site Investigator), Dick Trost, PhD (Cognitive Neurology - St. Joseph’s, Ontario, Site Investigator), Charles Bernick, MD (Cleveland Clinic Lou Ruvo Center for Brain Health, Site Investigator), Donna Munic, PhD (Cleveland Clinic Lou Ruvo Center for Brain Health, Site Investigator), Diana Kerwin, MD (Northwestern University, Site Investigator), Marek-Marsel Mesulam, MD (Northwestern University, Site Investigator), Kristina Lipowski, BA (Northwestern University, Site Investigator), Chuang-Kuo Wu, MD, PhD (Northwestern University, Past Site Investigator), Nancy Johnson, PhD (Northwestern University, Past Site Investigator), Carl Sadowsky, MD (Premiere Research Inst (Palm Beach Neurology), Site Investigator), Walter Martinez, MD (Premiere Research Inst (Palm Beach Neurology), Site Investigator), Teresa Villena, MD (Premiere Research Inst (Palm Beach Neurology), Site Investigator), Raymond Scott Turner, MD, PhD (Georgetown University Medical Center, Site Investigator), Kathleen Johnson, NP (Georgetown University Medical Center, Site Investigator), Brigid Reynolds, NP (Georgetown University Medical Center, Site Investigator), Reisa A. Sperling, MD (Brigham and Women’s Hospital, Site Investigator), Keith A. Johnson, MD (Brigham and Women’s Hospital, Site Investigator), Gad Marshall, MD (Brigham and Women’s Hospital, Past Site Investigator), Meghan Frey (Brigham and Women’s Hospital, Past Site Investigator), Jerome Yesavage, MD (Stanford University, Site Investigator), Joy L. Taylor, PhD (Stanford University, Site Investigator), Barton Lane, MD (Stanford University, Site Investigator), Allyson Rosen, PhD (Stanford University, Past Site Investigator), Jared Tinklenberg, MD (Stanford University, Past Site Investigator), Marwan Sabbagh, MD, FAAN, CCRI (Banner Sun Health Research Institute, Site Investigator), Christine Belden, PsyD (Banner Sun Health Research Institute, Site Investigator), Sandra Jacobson, MD (Banner Sun Health Research Institute, Site Investigator), Neil Kowall, MD (Boston University, Site Investigator), Ronald Killiany, PhD (Boston University, Site Investigator), Andrew E. Budson, MD (Boston University, Site Investigator), Alexander Norbash, MD (Boston University, Past Site Investigator), Patricia Lynn Johnson, BA (Boston University, Past Site Investigator), Thomas O. Obisesan, MD, MPH (Howard University, Site Investigator), Saba Wolday, MSc (Howard University, Site Investigator), Salome K. Bwayo, PharmD (Howard University, Past Site Investigator), Alan Lerner, MD (Case Western Reserve University, Site Investigator), Leon Hudson, MPH (Case Western Reserve University, Site Investigator), Paula Ogrocki, PhD (Case Western Reserve University, Site Investigator), Evan Fletcher, PhD (University of California, Davis – Sacramento, Site Investigator), Owen Carmichael, PhD (University of California, Davis – Sacramento, Site Investigator), John Olichney, MD (University of California, Davis – Sacramento, Site Investigator), Charles DeCarli, MD (University of California, Davis – Sacramento, Past Site Investigator), Smita Kittur, MD (Neurological Care of CNY, Site Investigator), Michael Borrie, MB ChB (Parkwood Hospital, Site Investigator), T-Y Lee, PhD (Parkwood Hospital, Site Investigator), Dr Rob Bartha, PhD (Parkwood Hospital, Site Investigator), Sterling Johnson, PhD (University of Wisconsin, Site Investigator), Sanjay Asthana, MD (University of Wisconsin, Site Investigator), Cynthia M. Carlsson, MD (University of Wisconsin, Site Investigator), Steven G. Potkin, MD (University of California, Irvine – BIC, Site Investigator), Adrian Preda, MD (University of California, Irvine – BIC, Site Investigator), Dana Nguyen, PhD (University of California, Irvine – BIC, Site Investigator), Pierre Tariot, MD (Banner Alzheimer’s Institute, Site Investigator), Adam Fleisher, MD (Banner Alzheimer’s Institute, Site Investigator), Stephanie Reeder, BA (Banner Alzheimer’s Institute, Site Investigator), Vernice Bates, MD (Dent Neurologic Institute, Site Investigator), Horacio Capote, MD (Dent Neurologic Institute, Site Investigator), Michelle Rainka, PharmD, CCRP (Dent Neurologic Institute, Site Investigator), Douglas W. Scharre, MD (Ohio State University, Site Investigator), Maria Kataki, MD, PhD (Ohio State University, Site Investigator), Earl A. Zimmerman, MD (Albany Medical College, Site Investigator), Dzintra Celmins, MD (Albany Medical College, Site Investigator), Alice D. Brown, FNP (Albany Medical College, Past Site Investigator), Godfrey D. Pearlson, MD (Hartford Hosp, Olin Neuropsychiatry Research Center, Site Investigator), Karen Blank, MD (Hartford Hosp, Olin Neuropsychiatry Research Center, Site Investigator), Karen Anderson, RN (Hartford Hosp, Olin Neuropsychiatry Research Center, Site Investigator), Andrew J. Saykin, PsyD (Dartmouth-Hitchcock Medical Center, Site Investigator), Robert B. Santulli, MD (Dartmouth-Hitchcock Medical Center, Site Investigator), Eben S. Schwartz, PhD (Dartmouth-Hitchcock Medical Center, Site Investigator), Kaycee M. Sink, MD, MAS (Wake Forest University Health Sciences, Site Investigator), Jeff D. Williamson, MD, MHS (Wake Forest University Health Sciences, Site Investigator), Pradeep Garg, PhD (Wake Forest University Health Sciences, Site Investigator), Franklin Watkins, MD (Wake Forest University Health Sciences, Past Site Investigator), Brian R. Ott, MD (Rhode Island Hospital, Site Investigator), Henry Querfurth, MD (Rhode Island Hospital, Site Investigator), Geoffrey Tremont, PhD (Rhode Island Hospital, Site Investigator), Stephen Salloway, MD, MS (Butler Hospital, Site Investigator), Paul Malloy, PhD (Butler Hospital, Site Investigator), Stephen Correia, PhD (Butler Hospital, Site Investigator), Howard J. Rosen, MD (UC San Francisco, Site Investigator), Bruce L. Miller, MD (UC San Francisco, Site Investigator), Jacobo Mintzer, MD, MBA (Medical University South Carolina, Site Investigator), Crystal Flynn Longmire, PhD (Medical University South Carolina, Site Investigator), Kenneth Spicer, MD, PhD (Medical University South Carolina, Site Investigator), Elizabether Finger, MD (St. Joseph’s Health Care, Site Investigator), Irina Rachinsky, MD (St. Joseph’s Health Care, Site Investigator), John Rogers, MD (St. Joseph’s Health Care, Site Investigator), Andrew Kertesz, MD (St. Joseph’s Health Care, Past Site Investigator), Dick Drost, MD (St. Joseph’s Health Care, Past Site Investigator), Nunzio Pomara, MD (Nathan Kline Institute, Site Investigator), Raymundo Hernando, MD (Nathan Kline Institute, Site Investigator), Antero Sarrael, MD (Nathan Kline Institute, Site Investigator), Susan K. Schultz, MD (University of Iowa, Site Investigator), Laura L. Boles Ponto, PhD (University of Iowa, Site Investigator), Hyungsub Shim, MD (University of Iowa, Site Investigator), Karen Elizabeth Smith, RN (University of Iowa, Site Investigator), Norman Relkin, MD, PhD (Cornell University), Gloria Chaing, MD (Cornell University), Lisa Raudin, PhD (Cornell University), Amanda Smith, MD (University of South Florida), Kristin Fargher, MD (University of South Florida), Balebail Ashok Raj, MD (University of South Florida).

